# Uptake and acceptability of cervical cancer screening among female sex workers in Eastern Uganda: A cross-sectional study

**DOI:** 10.1101/2024.10.18.24315734

**Authors:** Ronald Opito, Emmanuel Tiyo Ayikobua, Hellen Akurut, Susan Alwedo, Saadick Mugerwa Ssentongo, Walter Drake Erabu, Lazarus Oucul, Musa Kirya, Lameck Lumu Bukenya, Elly Ekwamu, Abraham Ignatius Oluka, Samuel Kabwigu, Emmanuel Othieno, Amos Deogratius Mwaka

**Author notes:** **Corresponding author:** Ronald Opito.

## Abstract

**Background:** Cervical cancer screening program in Uganda is opportunistic and focuses mainly on women aged 25-49 years. Female sex workers (FSWs) are at increased risk of developing invasive cervical cancer. There is limited data regarding the uptake and acceptability of cervical cancer screening among FSWs in Uganda. This study aimed at identifying factors affecting uptake and acceptability of cervical cancer screening among FSWs in Eastern Uganda.

**Methods:** This was a cross-sectional study conducted among 423 FSWs aged 18-49 years attending care at six health facilities serving Key Population in the Teso sub-region. Data was collected using structured investigator administered questionnaire and analyzed using Stata statistical software version 15.0 (Stata Corp, Texas, USA). The primary outcome was uptake of cervical cancer screening measured as the proportion of female sex workers who have ever been screened for cervical cancer. Chi-square test was used to compare the differences in uptake of cervical cancer screening by HIV status. Modified Poisson regression model with a robust variance estimator was used to determine association between the outcome variables and selected independent variables including demographic characteristics. Prevalence ratios (PR) with accompanying 95% confidence intervals have been reported. Statistical significance was considered at two-sided p-values ≤ 0.05.

**Results:** The mean age of the participants was 28.1(±SD=6.6) years. The self-reported HIV prevalence was 21.5% (n=91). There were 138 (32.6%) participants who had ever been screened for cervical cancer (uptake), while 397 (93.9%) were willing to be screened (acceptability). There was a significant difference in cervical cancer screening uptake between women living with HIV and those who were HIV negative, 59.3% vs 26.9% respectively (P<0.001). The significant factors associated with uptake of cervical cancer screening included being HIV positive, adjusted prevalence ratio (aPR) = 1.74; (95% CI: 1.32-2.29), living near a private not for profit (PNFP) facility, aPR = 2.02 (95% CI; 1.38-2.95), availability of screening services at the nearest health facility, aPR=1.66 (95% CI, 1.16-2.37) and being currently on pre-exposure prophylaxis (PrEP), aPR=1.62 (95% CI, 1.12-2.34). Factors significantly associated with acceptability included never screening for cervical cancer, aPR=1.08 (95%CI, 1.01-1.14), and living near a PNFP facility, aPR=1.12 (95% CI, 1.06-1.19).

**Conclusion:** Female sex workers living with HIV are more likely to screen for cervical cancer than the HIV negative clients. Cervical cancer screening uptake is relatively low among the female sex workers. However, majority of the FSWs are willing to be screened for cervical cancer if the services are provided in the nearby healthcare facilities. There is need to make cervical cancer screening services available to all eligible women especially the female sex workers and integrate the services with sexual reproductive health services in general and not just HIV/ART clinics services.

## Introduction

Cervical cancer is still a significant public health concern among females worldwide. In 2022, cervical cancer was the 8^th^ commonest cancer and 9^th^ cause of cancer death worldwide, with an estimated 661,021 women diagnosed with cervical cancer and 348,189 women that died from the disease [1]. Globally, the age standardized incidence rate of cervical cancer was 14.1 per 100,000 women [1]. However, in the low- and middle-income countries (LMICs) with low human development index (HDI), cervical cancer is still one of the commonest (second most common) cancers accounting for significant mortality. In the countries with high human development index, the incidence of and mortality from cervical cancer were 12.1 and 4.8 per 100,000 females, while in the countries with low HDI, the incidence and mortality were up to 19.3 and 12.4 per 100,000 females respectively [1]. In East Africa, the incidence and mortality of cervical cancer were as high as 40.4 and 28.9 per 100,000 women respectively, compared to corresponding figures of 6.6 and 2.1 per 100,000 women respectively in western Europe [1]. In Uganda, there were an estimated 6,938 new cervical cancer cases diagnosed, with 4,782 deaths from the disease in 2022, translating into age standardized incidence and mortality of 53.8 and 40.6 per 100,000 women respectively [2].

Cervical cancer is primarily caused by Human papillomavirus (HPV) 16 and 18, that contribute for about 70% of the global new cervical cancer cases [3]. Women living with HIV are at increased risk of cervical cancer as compared to their HIV negative counterparts [4–6]. Other factors associated with development of cervical cancer and HPV positivity include; early sexual debut [7], history of having a sexually transmitted infection [5,6,8] and having more than 2 lifetime sexual partners [6]. There is a disproportionately high prevalence of HPV and a correspondingly high prevalence of cervical cancer among female sex workers globally [7,8]. Female sex workers are at a high risk of acquiring HPV, HIV and other sexually transmitted infections (STIs) mainly because of having multiple sex partners and having unprotected sex, both of which are facilitators of cervical cancer development [9]. This subpopulation of women therefore requires increased attention to protect them from developing cervical cancer or at least screen them regularly to detect cancer in pre-invasive phase or early stage when treatment can be beneficial.

Cervical cancer can be eliminated and prevented by using proven cost effective measures including scheduled screening to detect early lesions that can be treated effectively [10]. The global strategy to accelerate the elimination of cervical cancer by 2030 involves the three pillars of achieving: 90% Human papillomavirus (HPV) vaccination coverage of eligible girls, 70% screening coverage with a high-performance test, and 90% of women with a positive screening test or a cervical lesion managed appropriately [3]. Cervical cancer screening in the HICs has generally led to the detection of more than 90% of all cancer cases before they metastasize to other parts of the body [11]. The success of screening depends on access and uptake, quality of screening tests, adequacy of follow-up, and prompt diagnosis and treatment of lesions detected [10]. However, coverage of cervical cancer screening services is low in most LMICs with an average estimated at 19% [12]. A community-based study in rural eastern Uganda showed a screening uptake of just 4.8% (43/900) [10]. Similarly, a community-based study in western Uganda showed low screening uptake of 7.0% (29/416) [13]. Even in circumstances where cervical screening services are available, uptake of screening is low due to several barriers. The barriers to cervical cancer screening include negative individual perceptions, n=553 (64.5%) and health facility related challenges, n=142 (16.6%)[10]. A study among HIV positive women in northern Uganda with a screening prevalence of 58.9% (175/279) showed that barriers to cervical cancer screening uptake include not being sure of benefits of screening and fear of screening outcomes while facilitators of screening included hearing about cervical cancer before study and knowing where cervical screening is conducted [15]. In western Uganda, the screening prevalence among HIV positive women was lower, 39.1% (161/412) compared to that in northern Uganda. The barriers to screening among the women in western Uganda included fear of positive screening results, and the perceptions that screening is expensive, embarrassing, painful and consumes a lot of time [16]. In general, a review of studies conducted in Uganda showed that barriers to cervical cancer screening in Uganda include limited access to healthcare facilities that offer screening, lack of resources to reach screening, perceived embarrassment and or pain from the procedure, fear of the screening procedure or outcome, and residing in a remote or rural area [17]. Similarly, a study in Cameroon also showed that barriers to cervical cancer screening include limited awareness of the benefits of screening and where screening is done, long distances to facilities that offer screening, high costs of screening, and negative attitudes and disrespect from the healthcare professionals that provide screening [18].

In Uganda, there are some data regarding uptake and acceptability of cervical screening in the general population. However, there is limited understanding of the factors affecting uptake and acceptability of cervical cancer screening among female sex workers in Uganda. Yet, female sex workers are particularly at high risk of developing and progressing to invasive cervical cancer [19]. This subpopulation of women also faces several barriers to cervical cancer screening, due to among others poor physical accessibility because of the highly mobile nature of their trade, they are a hard-to-reach population, and often the opening hours for clinics are not convenient for them [20,21]. This study aimed at determining the level of uptake, acceptability of cervical cancer screening, and associated factors among female sex workers in Northeastern Uganda.

## Materials and Methods

### Study Design and Setting

This was a cross-sectional study conducted in Teso sub-region in Northeastern Uganda. The Teso sub-region comprises 10 districts of Soroti, Amuria, Kapelebyong, Katakwi, Bukedea, Kumi, Ngora, Serere, Kalaki and Kaberamaido and Soroti City, with a total population of 1,819,708 [22]. The HIV prevalence in the region is 4.2% [23]. The total number of health facilities supported by The United States President’s Emergency Plan for AIDS Relief (PEPFAR) through Center for Disease Control and prevention (CDC) to provide high quality HIV and cervical cancer screening services is 106 (1 RRH, 7 Hospitals, 10 HCIV, 62 HCIII, 23 HCII and 3 Special clinics), of which 17 offer key and priority population services and had served 2043 female sex workers between October 2021 and March 2022 (https://mets.or.ug/kp-dashboard). These health facilities are categorized and ranked by levels, with Health center III (HCIII) being the lowest supported health facility and regional referral hospital (RRH) being the highest tertiary health facility supported and special clinics being those health facilities specialized in the provision of HIV prevention, care, and treatment services. The estimated population of female sex workers in the region is 5784 [24]. The central point of the region is Soroti district which is located 326 km Northeast of Uganda’s capital Kampala. Our study purposively selected six (6) high volume health facilities which served at least 100 female sex workers in six months. These included Katakwi hospital, Toroma HCIV, Kumi HCIV, Eastern division HCIII, Kichinjajji HCIII and TASO Soroti Centre of Excellence, which in total had served 1633 (80%) female sex workers in the region.

### Study Population

The study population included all female sex workers 18-49 years of age receiving HIV prevention, care, and treatment services in the Teso sub-region during the study period. We included all female sex workers, who were willing and able to provide voluntary informed consents in the study. We excluded female sex workers who had not yet attained majority age, women with a history of hysterectomy and those who were very ill at the time of data collection.

### Sample size estimation

The sample size was estimated using the Kish Leslie equation (1965) [25]. To get large sample size that can help evaluate several factors associated with uptake, we used uptake prevalence of 50% since there are very scarce study in this subpopulation. We obtained a sample size of 384 and adjusted for 10% non-response to obtain a sample size of 422 participants. In our final analysis, we included 423 participants distributed in the six facilities proportionate to the number of individuals served per facility as follows: Katakwi Hospital (40), Toroma HCIV (27), Kumi HCIV (200), Eastern Division HCIV (30), Kichinjajji HCIII (93) and TASO Soroti Centre of Excellence (33).

### Participants sampling and sampling technique

We used consecutive sampling to recruit the female sex workers coming for HIV prevention and treatment services and willing to participate in the study. We consented and recruited all those who accepted to participate in the study. Recruitment and data collection were conducted during March 2023. Prospective participants were recruited until each site sample size was reached.

### Study Variables

The primary outcome variable was uptake of cervical cancer screening among female sex workers. Cervical cancer screening uptake was measured as self-reported ever undertaking of a cervical cancer screening test. Respondents were specifically asked “Have you ever been tested or examined for cervical cancer or precancer?” (No/Yes). The secondary outcome variable was acceptability of cervical cancer screening where participants were asked whether they were willing to accept cervical cancer screening if provided with the opportunity (No/Yes). The independent variables included socio-demographic information including age, religion, occupations, place of residence, ethnicity, family history of cervical cancer, number of sexual partners, marital status, and parity. We also included other health characteristics based on findings from previous studies. These factors included self-reported use of family planning, HIV status, history of STI, and recent visit to health facility.

### Data collection and management

Data was collected using investigator administered questionnaire installed into android phones loaded with the online Kobo toolbox software (https://www.kobotoolbox.org/). We trained ten (10) research assistants (5 female and 5 male) who were medical/nursing students at Soroti University to recruit and interview participants. The research assistants (RA) were trained on the study objectives, matters related to the female sex work, and confidentiality required during research, consenting process, and the interview technique using the kobo toolbox for online data collection. They were also trained on aspects of cervical cancer including the burden, risk factors and symptoms, and the different approaches to prevention, screening and early detection approaches, and management. The RAs approached the clients when they had come to receive clinical care including HIV prevention education, care for sexually transmitted infections (STI), and treatment for other diseases. The RAs explained the purpose and process of the study to the prospective participants. Prospective participants were allowed to ask questions regarding the study. After their concerns were addressed, the study participants were offered and completed the informed consent forms before interviews. We administered the questionnaire to all eligible study participants that provided informed consent. Figure 1 below shows the recruitment flow chart.

**Figure1:**
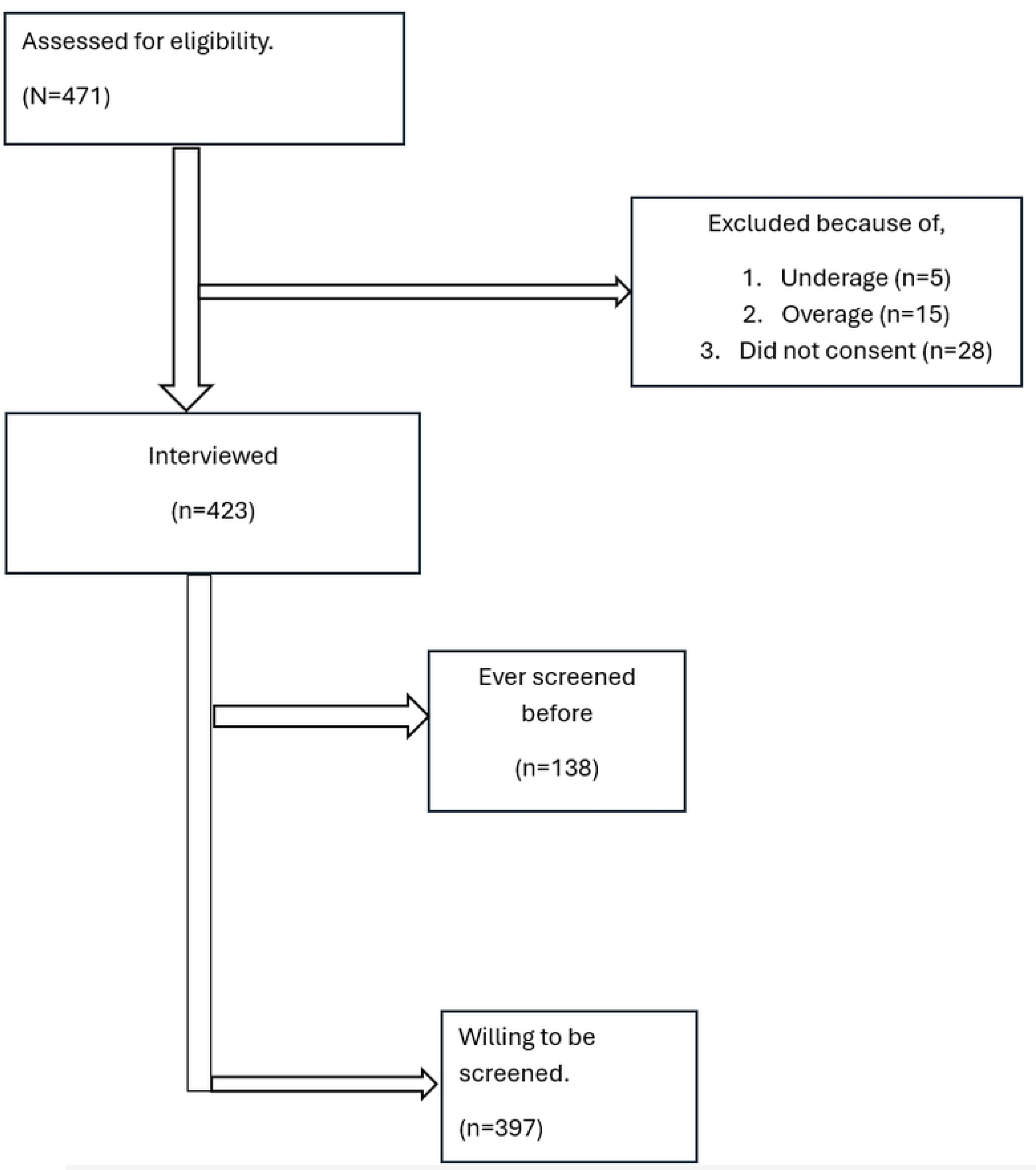
Participant Recruitment flow diagram

### Data Analysis

Data from the Kobo toolbox software was downloaded onto an excel sheet at the end of each day of data collection. RO and AIO reviewed the data for completeness after download and before storage. Incomplete records were completed with help of the RAs and or by calling the participants. Those that could not be completed reliably were removed from the dataset. Duplicated participant unique ID were corrected to ensure each unique ID is assigned to only one participant. RO and AIO cleaned the data, assessed for missing values, and inconsistent entries. The cleaned data was then exported to STATA version 15.0 statistical software (Stata Corp, Texas, USA) for further management including analysis. RO and AIO conducted data analysis. At univariate analysis, the socio-demographics including age, marital status, and duration of sex work was summarized and presented as frequencies, median with interquartile ranges, and mean with standard deviations. Both the primary outcome, which was uptake of cervical cancer screening, and the secondary outcome, acceptability of cervical cancer screening was presented as proportions. We conducted bivariate analysis between the pre-selected independent predictor variables mentioned above and the two outcome variables. We then used Modified Poisson regression model with a robust variance estimator to determine factors associated with uptake and acceptability of cervical cancer screening among FSWs. Modified Poisson was chosen over logistic regression because the outcome is common (based on similar studies), and we wanted to estimate prevalence ratios instead of odds ratios. Before conducting the multivariate Poisson regression, the pre-selected variables were included in a correlation model and checked for multicollinearity. Where variables showed multicollinearity (r ≥ 0.4), only one was included in the final model. Each of the pre-selected independent/predictor variable was included into separate independent models, and was adjusted against the sociodemographic characteristics - age, parity, marital status, education level, employment, HIV status and family history of cervical cancer. The measures of effect sizes reported are prevalence ratios with their corresponding 95% confidence intervals. Variables with two-sided p-value < 0.05 were considered statistically significant.

### Ethical Approval and consent to participate

Ethical clearance for this study was sought from the TASO Research and Ethics Committee (REC), approval number, TASO-2022-151. Administrative clearances were sought from the office of the district health officer (DHO) of the respective districts and permission to conduct the study sought from the in-charges of the facilities where the study was conducted. The individual participant provided written informed consent after the research assistants had explained to them the aims of the study, the benefits and risks associated with the study. Participants’ information was anonymized before entry into the database for analysis. This was to ensure confidentiality and privacy. We reported the results as disaggregated data without revealing individual information.

## Results

### Socio-demographic characteristics

The mean age of the 423 participants was 28.2 (SD=6.6) years; the majority were young, aged 20 –29 years (54.1%, n=229), never married (62.9%, n=266), and with primary level education (44.2%, n=187). Most participants (70.9%, n=300) had been in sex work for less than five years, and were current users of hormonal contraceptives (82.5%, n=349). Majority had no history of previous sexually transmitted infections (61.8%, n=257). The self-reported HIV prevalence in this study was 21.5% (n=91). Nearly half of the HIV negative participants (42.6%, n=130) were on pre-exposure prophylaxis. Most participants (94.6%, n=400) lived within less than five kilometers to the nearest health facility (Table 1).

**Table 1:** Socio-demographic and health related characteristics of study participants N=423.

| **Factors** | **Frequency (n)** | **Proportions (%)** |
| --- | --- | --- |
| **Continuous variables** |  |  |
| Current age (Years) | 28.2 (6.6) |  |
| **Age Category** |  |  |
| <20 Years | 30 | 9.1 |
| 20-29 Years | 229 | 54.1 |
| 30-39 Years | 142 | 33.6 |
| 40+ Years | 22 | 5.2 |
| **Marital status** |  |  |
| Never married (single) | 266 | 62.9 |
| Married | 18 | 4.3 |
| Widow | 18 | 4.3 |
| Divorced/separated | 121 | 28.6 |
| **Duration of sex work** |  |  |
| <5years | 300 | 70.9 |
| ≥5years | 123 | 29.1 |
| **Parity** |  |  |
| <3children | 247 | 58.4 |
| ≥3children | 176 | 41.6 |
| **Education level** |  |  |
| No formal education | 40 | 9.5 |
| Primary level | 187 | 44.2 |
| Secondary Level | 165 | 39.0 |
| Tertiary level | 31 | 7.3 |
| **Formal employment** |  |  |
| Unemployed | 193 | 45.6 |
| Employed | 230 | 54.4 |
| **Cervical cancer screening services is available in the nearest health facility to my residence** |  |  |
| Yes | 289 | 68.3 |
| No/Not sure | 134 | 31.7 |
| **Distance to nearest health facility (kilo meters)** |  |  |
| <5km | 400 | 94.6 |
| ≥5km | 23 | 5.4 |
| **Self-reported HIV status** |  |  |
| Negative | 305 | 72.1 |
| Positive | 91 | 21.5 |
| Unknown | 27 | 6.4 |
| **Current PrEP User for HIV negative (N=305)** |  |  |
| No | 175 | 57.4 |
| Yes | 130 | 42.6 |
| **Using hormonal family planning during study time** |  |  |
| No | 74 | 17.5 |
| Yes | 349 | 82.5 |
| **Ever been diagnosed with STI in recent one year** |  |  |
| No | 257 | 61.8 |
| Yes | 166 | 38.2 |
| **Age at first sex** |  |  |
| <15Years | 105 | 24.8 |
| ≥15years | 318 | 75.2 |
| **Family history of cervical cancer** |  |  |
| No | 339 | 80.1 |
| Yes | 84 | 19.9 |

STI = sexually transmitted infections; PrEP = Pre-exposure prophylaxis.

### Socio-demographic factors associated with cervical cancer screening uptake

Cervical cancer screening uptake was 32.6% (95%CI; 28.3 -37.3), (n=138). There was a significant difference in cervical cancer screening uptake between women living with HIV and those who were HIV negative, 59.3% vs 26.9% (P<0.001) respectively. At multivariate level, self-reported cervical cancer screening was independently associated with being HIV positive, adjusted prevalence ratio, aPR = 1.74 (95%CI; 1.32-2.29) (Table 2).

**Table 2:** Socio-demographic factors associated with uptake of cervical cancer screening.

| Characteristic | Cervical cancer screening status |  | Prevalence ratio (PR) |  |
| --- | --- | --- | --- | --- |
|  | Ever screened.<br>n (%),<br>138 (32.6) | Never Screened<br>n (%),<br>285 (67.4) | Crude PR<br>(95%CI) | Adjusted PR<br>(95%CI) |
| **Age (Years)** |  |  |  |  |
| <20 | 4 (13.3) | 26 (86.7) | 1 | 1 |
| 20-29 | 58 (25.3) | 171 (74.7) | 1.90(0.74-4.86) | 1.38(0.54-3.53) |
| 30-39 | 65 (45.8) | 77 (54.2) | **3.43(1.35-8.71)** | 1.81(0.68-4.83) |
| ≥ 40 | 11 (50.0) | 11 (50.0) | **3.75(1.37-10.24)** | 1.61(0.55 - 4.70) |
| **Marital Status** |  |  |  |  |
| Never married | 72 (27.1) | 194 (72.9) | 1 | 1 |
| Married | 5 (27.9) | 13 (72.2) | 1.02(0.47-2.22) | 0.86(0.43-1.72) |
| Widowed | 12 (66.7) | 6 (33.3) | **2.46(1.68-3.61)** | 1.33(0.88-2.00) |
| Divorced/Separated | 49 (40.5) | 72 (59.5) | **1.50(1.12-2.01)** | 1.16(0.85-1.58) |
| **Parity** |  |  |  |  |
| <3 Children | 62 (25.1) | 185 (74.9) | 1 | 1 |
| ≥3 Children | 76 (43.2) | 100 (56.8) | **1.72 (1.31-2.26)** | 1.27(0.94-1.71) |
| **Educational attainment** |  |  |  |  |
| No formal education | 19 (47.5) | 21 (52.5) | 1 | 1 |
| Primary | 63 (33.7) | 124 (66.3) | 0.71(0.48-1.04) | 0.93(0.63-1.38) |
| Secondary | 38 (23.0) | 127 (77.0) | **0.48(0.32-0.74)** | 0.70(0.44-1.10) |
| Tertiary | 18 (58.1) | 13 (41.9) | 1.22(0.79-1.90) | 1.60(0.98-2.60) |
| **Formal employment Status** |  |  |  |  |
| Unemployed | 69 (35.7) | 124 (64.3) | 1 | 1 |
| Employed | 69 (30.0) | 161 (70.0) | 0.84(0.64-1.10) | 0.87(0.67-1.15) |
| **HIV status** |  |  |  |  |
| Negative | 82 (26.9) | 223 (73.1) | 1 | 1 |
| Positive | 54 (59.3) | 37 (40.7) | **2.21(1.71-2.84)** | **1.74(1.32-2.29)** |
| **Ever been diagnosed with STI** |  |  |  |  |
| No | 84(32.7) | 173 (67.3) | 1 | 1 |
| Yes | 54 (32.5) | 112 (67.5) | 1.00(0.75-1.32) | 0.97(0.74-1.27) |
| **There is a family history of cervical cancer** |  |  |  |  |
| No | 109 (32.1) | 230 (67.9) | 1 | 1 |
| Yes | 29 (34.5) | 55 (65.5) | 1.07 (0.77-1.50) | 1.05 (0.78-1.41) |
| **Duration in sex work (years)** |  |  |  |  |
| <5 | 91 (30.3) | 209 (69.7) | 1 | 1 |
| ≥5 | 47 (38.2) | 76 (61.8) | 1.26(0.95-1.67) | 1.00 (0.74-1.36) |

**Bold** = Significant with $P < 0.05$ ; All factors were adjusted for each other.

### Health system factors associated with cervical cancer screening uptake

Participants who lived near private not for profit (PNFP) health facilities were twice as likely to have undergone cervical cancer screening than those who lived nearest a public (government owned) facility, aPR = 2.02 (95%CI; 1.38 - 2.95). Regarding level of health facility, participants living near to a health center IV were less likely to have undergone cervical cancer screening than those living near to a health center III, aPR = 0.67 (95% CI; 0.49 - 0.93). Availability of cervical cancer screening services in the nearest health facility was statistically significantly associated with uptake of cervical cancer, aPR=1.66 (95% CI; 1.16 - 2.37). Female sex workers who were currently receiving PrEP were more likely to have undergone cervical cancer screening, aPR=1.62 (95%CI, 1.12 - 2.34) (Table 3).

**Table 3:**
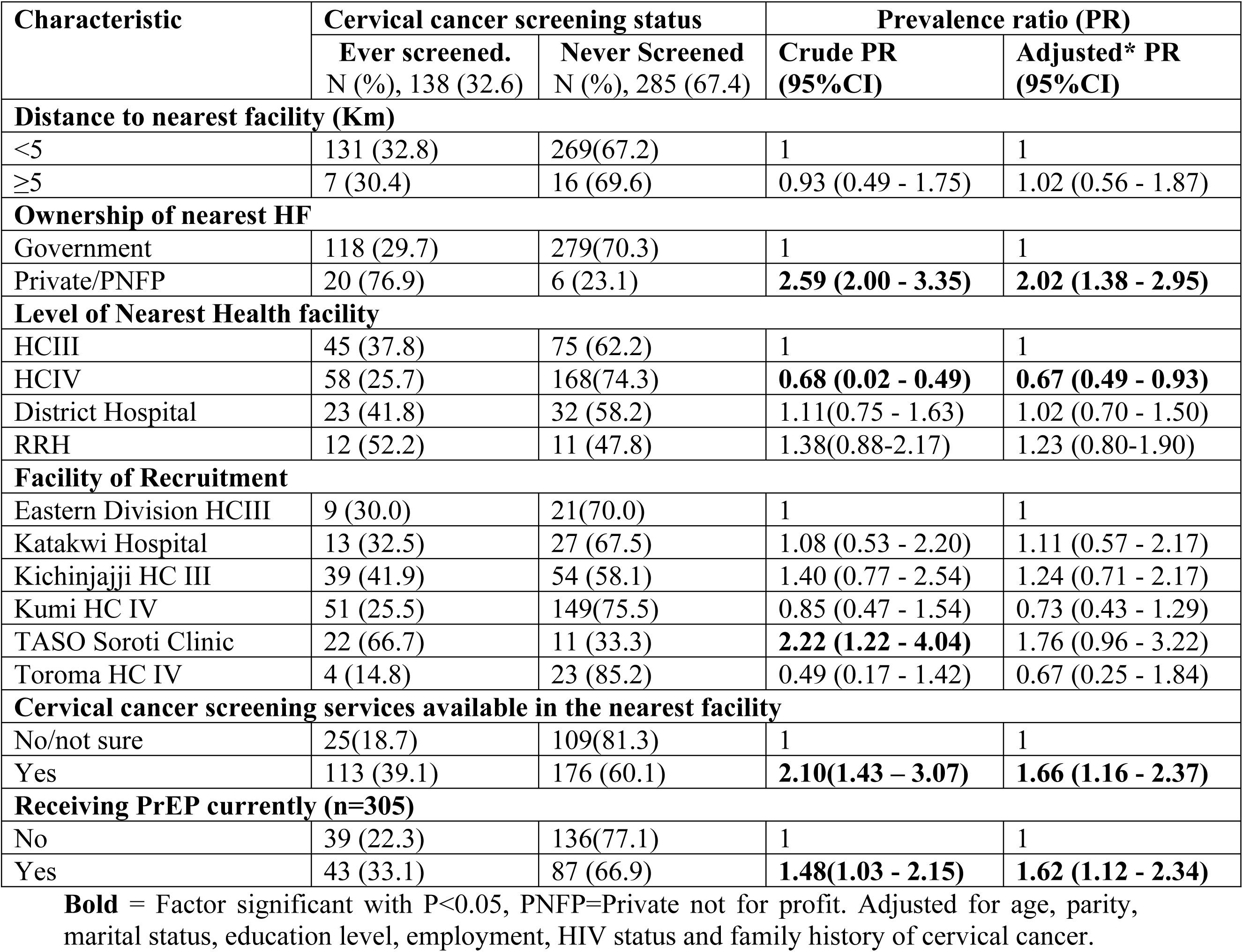
Health System factors and uptake of cervical cancer screening.

### Willingness to undertake cervical cancer screening (Acceptability)

Most participants, 397 (93.9%; 95%CI, 91.1 - 95.8) would accept cervical cancer screening if offered. On adjusting the sociodemographic variables for each other, acceptability of cervical screening was not statistically associated with any demographic factors (Table 4).

**Table 4.** Sociodemographic factors associated with willingness to undertake cervical cancer screening (acceptability) Bold = Factor significant with P<0.05. All factors adjusted for each other.

| Characteristic | Willingness to undertake cervical cancer screening |  | Prevalence Ratio (PR) |  |
| --- | --- | --- | --- | --- |
|  | Yes (*n*=397)<br>N (%) | No (*n*=26)<br>N (%) | Crude PR (95%CI) | Adjusted PR (95%CI) |
| **Age Category** |  |  |  |  |
| <20 Years | 25(83.3) | 5(16.7) | 1 | 1 |
| 20-29 Years | 218(95.2) | 11(4.8) | 1.14(0.97-1.34) | 1.18(0.95-1.47) |
| ≥30Years | 154(93.9) | 10(6.1) | 1.13(0.96-1.33) | 1.18(0.95-1.47) |
| **Marital status** |  |  |  |  |
| Never married | 249(93.6) | 17(6.4) | 1 | 1 |
| Ever married | 148(94.3) | 9(5.7) | 1.01(0.96-1.06) | 1.00(0.95-1.05) |
| **Parity** |  |  |  |  |
| <3children | 232(93.9) | 15(6.1) | 1 | 1 |
| ≥3children | 165(93.7) | 11(6.3) | 1.00(0.95-1.05) | 0.98(0.93-1.03) |
| **Education level** |  |  |  |  |
| No/primary | 210(92.5) | 17(7.5) | 1 | 1 |
| Post-primary | 187(95.4) | 9(4.6) | 1.03(0.98-1.08) | 1.02(0.97-1.08) |
| **Formal employment** |  |  |  |  |
| Unemployed | 176(91.2) | 17(8.8) | 1 | 1 |
| Employed | 221(96.1) | 9(3.9) | **1.05(1.00-1.11)** | 1.03(0.98-1.09) |
| **Duration of Sex work** |  |  |  |  |
| <5years | 280(93.3) | 20(6.7) | 1 | 1 |
| ≥5years | 117(95.1) | 6(4.9) | 1.02(0.97-1.07) | 1.02(0.96-1.08) |
| **HIV status** |  |  |  |  |
| Negative | 289(94.8) | 16(5.2) | 1 | 1 |
| Positive | 82(90.1) | 9(9.9) | 0.95(0.88-1.02) | 0.95(0.88-1.02) |
| **Family history of cervical cancer** |  |  |  |  |
| No | 315(92.9) | 24(7.1) | 1 | 1 |
| Yes | 82(97.6) | 2(2.4) | **1.05(1.00-1.10)** | 1.05(1.00-1.10) |

Participants who had never screened for cervical cancer were statistically more likely to accept future cervical screening compared to those who had ever screened, aPR=1.08(95%CI: 1.01 - 1.14), and participants living near to private not for profit (PNFP) health facilities were more likely to accept cervical cancer screening compared to those who lived nearest a public (government owned) facility, aPR=1.12 (95%CI: 1.06 - 1.19) (Table 5).

**Table 5:**
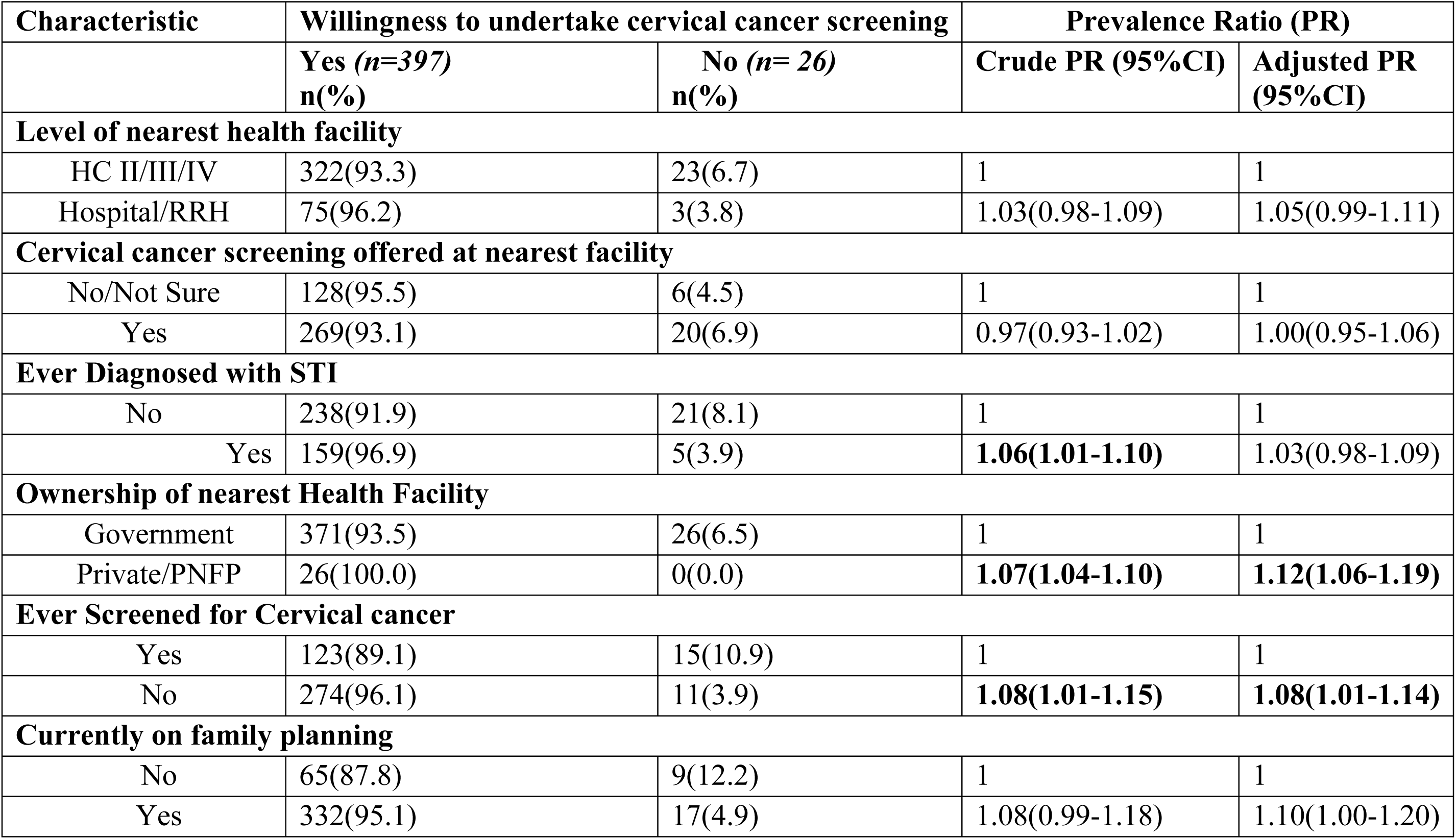
Health system factors and acceptability of cervical cancer screening. Bold=Significant with P<0.05. Adjusted for age, parity, marital status, education level, employment, HIV status, duration of sex work and family history of cervical cancer.

| Characteristic | Willingness to undertake cervical cancer screening |  | Prevalence Ratio (PR) |  |
| --- | --- | --- | --- | --- |
|  | Yes (*n*=397)<br>n(%) | No (*n*= 26)<br>n(%) | Crude PR (95%CI) | Adjusted PR (95%CI) |
| **Level of nearest health facility** |  |  |  |  |
| HC II/III/IV | 322(93.3) | 23(6.7) | 1 | 1 |
| Hospital/RRH | 75(96.2) | 3(3.8) | 1.03(0.98-1.09) | 1.05(0.99-1.11) |
| **Cervical cancer screening offered at nearest facility** |  |  |  |  |
| No/Not Sure | 128(95.5) | 6(4.5) | 1 | 1 |
| Yes | 269(93.1) | 20(6.9) | 0.97(0.93-1.02) | 1.00(0.95-1.06) |
| **Ever Diagnosed with STI** |  |  |  |  |
| No | 238(91.9) | 21(8.1) | 1 | 1 |
| Yes | 159(96.9) | 5(3.9) | **1.06(1.01-1.10)** | 1.03(0.98-1.09) |
| **Ownership of nearest Health Facility** |  |  |  |  |
| Government | 371(93.5) | 26(6.5) | 1 | 1 |
| Private/PNFP | 26(100.0) | 0(0.0) | **1.07(1.04-1.10)** | **1.12(1.06-1.19)** |
| **Ever Screened for Cervical cancer** |  |  |  |  |
| Yes | 123(89.1) | 15(10.9) | 1 | 1 |
| No | 274(96.1) | 11(3.9) | **1.08(1.01-1.15)** | **1.08(1.01-1.14)** |
| **Currently on family planning** |  |  |  |  |
| No | 65(87.8) | 9(12.2) | 1 | 1 |
| Yes | 332(95.1) | 17(4.9) | 1.08(0.99-1.18) | 1.10(1.00-1.20) |

## Discussion

In this study, most participants were young, attained low levels of formal education, and did not have formal employment. Most of the participants had never been married or ever got married but divorced/separated. The self-reported prevalence of cervical cancer screening (uptake) was relatively high though still lower than the WHO recommended prevalence of 70% [26]. The acceptability of cervical cancer screening (intention-to-use cervical screening services when availed) was high among the participants. Factors that were statistically significantly associated with uptake of cervical cancer screening included being HIV positive, living near to a private not for profit (PNFP) health facility, availability of cervical cancer screening services in the nearest health facility, and being currently on PrEP. The factors statistically significantly associated with acceptability were having not been screened previously and living near to a PNFP health facility.

There were prevalences of risk factors for development of cervical cancer in this study population. For example, the prevalence of self-reported HIV positivity was higher than the regional and national prevalence of 4.2% and 5.8% respectively[23]. The high prevalence of HIV infection among the sex workers in this study is not an isolated finding. HIV prevalence among female sex workers in Uganda is generally higher (33%) than HIV prevalence in the general population [27]. HIV infection is a risk factor for the development and progression of cervical cancer [4–6]. Similarly, the self-reported history of treatment for sexually transmitted infections (STIs) was high among this study participants. And there is evidence that sexually transmitted infections (STIs) including Chlamydia, Genital warts, Syphilis and Gonorrhea are associated with the development of cervical cancer [5,6,8]. In this study, a substantial proportion of participants (25%; n=105) reported having started penetrative vaginal sexual intercourse at age < 15 years, while 82.5% of the participants were using hormonal contraceptives. Early sexual debut is a known risk factor for invasive cervical cancer. At such early age, the ability of the immature cervical cells to resist dysplastic changes is still low, and this increases the risks of such women to developing cervical cancer in the future [7,28]. Long term use of hormonal contraceptives especially estrogen containing contraceptives has been found to substantially increase the risk of developing cervical cancer [29]. The female sex workers in this study region, and perhaps other regions of Uganda are exposed to risk factors that may lead to the development of invasive cervical cancer. They therefore need to be encouraged to adopt healthier lifestyles including increased use of pre-exposure prophylaxis to avoid contracting HIV, and regular cervical cancer screening.

Cervical cancer screening uptake by the female sex workers in this region is generally higher compared to findings from other studies conducted in the general communities in Uganda; prevalence of cervical screening uptake range was 4.8% in eastern region [10], 7.0% in western region[13], and 17.1% to 20.0% in central region [14,30,31]. Generally, cervical cancer screening uptake is lower in the general community in most countries in sub–Saharan Africa. For example, low cervical cancer screening uptake of 12.9% (95% CI: 10.20, 15.54) was reported in a review of 29 studies on cervical cancer screening uptake in sub–Saharan Africa [32]. The prevalence of cervical cancer screening uptake is higher among HIV positive women attending specialized HIV care compared to prevalence of screening uptake among women in the general community. A review of studies among women with HIV attending specialized HIV care in Uganda showed that cervical cancer screening prevalence ranged between 30% and 59% [15,16]. These findings are comparable to our findings perhaps because the health facilities where we conducted our study are among the facilities supported by TASO to provide specialized HIV prevention, care and cervical cancer screening in the Teso subregion. The facilities also offer health education to clients on matters such as benefits of cervical screening and use of prophylactic ART by female sex workers. The availability of these services in these facilities explains in part the relatively higher cervical cancer screening prevalence in our study. Cervical cancer screening uptake prevalence in our study is however lower than uptake in similar populations of female sex workers elsewhere. In Kenya, a study among 418 female sex workers showed that uptake of cervical cancer screening was 72.8% (n=305)[33]. Similarly, the prevalence of cervical cancer screening uptake was higher among female sex workers (38.6%; 236/611) in Vancouver, Canada [34]. However, the screening uptake in our study was higher than in a study among 230 sex workers in Ethiopia where the cervical cancer screening uptake prevalence was 13.3% [35]. Female sex workers are a high-risk population that needs deliberate interventions to reduce the incidence and mortality from cervical cancer. These interventions include awareness campaigns to promote behavioral changes such as stopping smoking and adopting cervical cancer screening to reduce their risk of developing the cancer as well as interventions to promote early detection of invasive cervical cancer. Increasing cervical cancer awareness and knowledge about cervical cancer risk factors and symptoms increases screening uptake by about five times (OR: 4.81; 95% CI: 3.06, 7.54) [32]. In addition, interventions to increase availability and physical access to health facilities that offer cervical screening services are critical in increasing uptake of cervical screening. In this study, participants nearer to the private not for profit (PNFPs) health facilities that provided cervical cancer screening services were more likely to have been screened. In other studies in Uganda and Jordan, the use of private health care services was a significant positive predictor of cervical cancer screening [36,37]. A scoping review of 19 studies conducted in Africa to determine interventions to increase cervical cancer screening uptake showed that most women were willing to screen before and after the interventions[38]. Lack of cervical screening services is potentially a barrier to cervical cancer screening uptake in sub–Saharan Africa.

There was high acceptability (94%) of cervical cancer screening among this study population. Participants who had never screened for cervical cancer and those living near a PNFP were statistically significantly more likely to accept cervical cancer screening if offered. Our finding of high cervical cancer screening acceptability is similar to earlier findings from Mbarara regional referral hospital (65%) in western Uganda [39] and Luwero HCIV (91%) in central Ugandan [40]. However, cervical cancer screening acceptability has varied significantly across regions; in west Africa, studies in Nigeria showed 48% acceptance [41] while in Ghana, acceptance was 82% [42]. The high prevalence of cervical screening acceptability among female sex workers is reassuring. It is therefore appropriate that the Ministry of health of Uganda leverages on the women’s willingness to screen and scale up cervical cancer screening especially in the high-risk population of female sex workers to promote early detection and prompt adequate treatment so as to minimize morbidity and mortality from cervical cancer.

### Limitations of the study

Our study had some limitations. The study adopted a quantitative survey design that could not lend itself to exploring the nuances regarding the reasons and motivations for the observed cervical screening uptake and acceptability among female sex workers. Such reasons that are necessary to inform design of targeted health interventions to promote cervical cancer screening among female sex workers could best be obtained from exploratory in-depth interviews and or focus group qualitative studies. Second, we based our ascertainment of outcome measures of uptake and acceptability as well as HIV infection status on participants reports which is susceptible to social desirability bias. Third, our findings ought to be interpreted with cautions since we used consecutive sampling approach to recruit clients into the study; this sampling approach potentially introduces selection and temporal biases. The participants that attended care during the period of the study recruitment can be significantly different in important ways from those that attend in other periods. These biases potentially reduce on representativeness and generalizability of the study findings. However, we are not aware of any unique background interventions or influences that could have differently and systematically driven the sex workers who attended care during the study period. These six health facilities are of varying levels of service provision (lower lever health facilities-HCIIIs, HCIVs, a specialized clinic and a higher-level facility, District hospital). This therefore provided a mix of participants and potentially countered the effects of selection and temporal bias which could have resulted from consecutive sampling.

## Conclusion

Female sex workers are aware of the benefits of screening for cervical cancer and are willing to be screened. However, availability of cervical cancer screening services is limited. The Uganda Ministry of Health may leverage on the women’s willingness to take up cervical cancer screening to scale up the service to promote identification of precancerous lesion in the high-risk population of sex workers, as well as enable early detection of invasive cervical cancer. These will contribute to prevention of invasive cervical cancer and allow for prompt effective treatment of early-stage invasive cervical cancer when treatments can still be effective. Integration of cervical cancer screening and treatment of precancerous lesions with the already established HIV care services is a feasible and sustainable approach to scale up cervical cancer screening and treatment for eligible women especially the female sex workers. There is also need for targeted health education to the female sex workers to promote health lifestyles including avoiding use of alcohol and cigarettes and use of pre-exposure prophylaxis in order to reduce their risks to development of invasive cervical cancer and contracting HIV respectively.

## Abbreviations

AIDS: Acquired immunodeficiency syndrome
CaCx: Cervical cancer
CDC: Centre for Disease Control and Prevention
FSWs: Female Sex Workers
HIV: Human Immunodeficiency virus
HPV: Human Papilloma Virus
KP: Key Populations
REC: Research Ethics Committee
TASO ’: The AIDS Support Organization
VIA: Visual Inspection with acetic acid
RRH: Regional Referral Hospital
HC: Health Centre
PEPFAR: President’s Emergency Plan for AIDS Relief.

## Availability of data and materials

The datasets used and/or analyzed during the current study are all included in this manuscript and its supporting information.

## Authors Contributions

**RO**: Conceptualization, proposal development, Methodology, funds acquisition and management, data collection, data analysis and report writing. **SA**, **LLB**, **LO:** Conceptualization, proposal development, Methodology and report writing. **SMS, HA, ETA, MK, EE:** Proposal development, methodology, data collection and report writing. **EO, SK, WDE:** Supervision, review and report writing. **AIO**: Data collection and data analysis. **ADM**: Review, methodology, data analysis, supervision and report writing.

## Data Availability

All relevant data are within the manuscript and its Supporting Information files.

## Acknowledgement.

The authors acknowledge the contribution of Professor JR Odong Ikoja, the Soroti University Vice Chancellor who acquired the funds for the research activities and Professor Francis Ejobi, the Soroti University Director of Research and Innovation for his administrative support and oversight on the management of this research project.

